# Leveraging Machine Learning for Enhanced and Interpretable Risk Prediction of Venous Thromboembolism in Acute Ischemic Stroke Care

**DOI:** 10.1101/2024.04.11.24305689

**Authors:** Youli Jiang, Ao Li, Zhihuan Li, Yanfeng Li, Rong Li, Qingshi Zhao, Guisu Li

## Abstract

**Background:** Stroke is the second leading cause of death globally, with acute ischemic strokes constituting the majority. Venous thromboembolism (VTE) poses a significant risk during the acute phase post-stroke, and early recognition is critical for preventive intervention of VTE.

**Methods:** Utilizing data from the Shenzhen Neurological Disease System Platform to develop multiple machine learning models that included variables such as demographics, clinical data, and laboratory results. Advanced technologies such as K nearest neighbor and synthetic minority oversampling technique are used for data preprocessing, and algorithms such as gradient boosting machine and support vector machine are used for model development.Feature analysis of optimal models using SHapley Additive exPlanations interpretable algorithm.

**Results:** In our study of 1,632 participants, in which women were more prevalent, the median age of patients with VTE was significantly older than that of non-VTE individuals. Data analysis showed that key predictors such as age, alcohol consumption, and specific medical conditions were significantly associated with VTE outcomes. The AUC of all prediction models is above 0.7, and the GBM model shows the highest prediction accuracy with an AUC of 0.923. These results validate the effectiveness of this model in identifying high-risk patients and demonstrate its potential for clinical application in post-stroke VTE risk management.

**Conclusion:** This study presents an innovative, machine learning-based approach to predict VTE risk in acute ischemic stroke patients, offering a tool for personalized patient care. Future research could explore integration into clinical decision systems for broader application.

## Introduction

Stroke remains a paramount health challenge worldwide, ranking as the second leading cause of mortality and contributing to approximately 6.1 million deaths each year. It also stands as a principal cause of long-term disability(1). Among its subtypes, acute ischemic strokes (AIS) predominate, significantly influencing the convalescence and functional recovery of patients. A critical concern in the aftermath of an ischemic stroke is the development of venous thromboembolism (VTE), a severe complication typically arising within the first two weeks post-event, with its highest risk noted in the initial seven days(2, 3). The occurrence of VTE is closely linked with an escalated risk of mortality within three months following a stroke, highlighting its importance as a preventable aspect of post-stroke care management(4, 5).

The arena of VTE risk prediction in acute ischemic stroke patients is riddled with complexities. This is largely due to the dependence on traditional risk assessment models such as the Caprini scoring system and the Padua Prediction Score, which inadequately address the multifaceted risk factor interplay inherent to this patient group. These models often overlook critical elements such as demographic nuances, clinical manifestations, and the detailed medical histories specific to individuals who have suffered a stroke(6–8). Given the heterogeneous nature of stroke survivors, accurately predicting VTE risk necessitates consideration of a myriad of determinants, including age, gender, pharmacotherapy, and chronic conditions like hypertension and diabetes, which are pivotal in assessing VTE risk(9–11). Acknowledging these challenges, our research introduces an innovative machine learning-based predictive model tailored for acute ischemic stroke patients. This model transcends traditional assessment tools by integrating an expansive range of risk factors—from demographic specifics to comprehensive clinical and laboratory data—leveraging advanced algorithms to enhance interpretability. Our approach not only aims to achieve superior accuracy in VTE risk prediction but also seeks to shed light on the relative importance of various risk factors, thereby offering a more nuanced and personalized risk assessment for stroke survivors(12).

Furthermore, the literature reveals a conspicuous scarcity of studies employing advanced machine learning techniques in conjunction with SHapley Additive exPlanations (SHAP) interpretability algorithms to forecast VTE subsequent to AIS. There exists a notable gap in the research landscape concerning the development of machine learning models that not only predict VTE with high precision but also adhere to the stringent criteria set forth by the Predictive Model Bias Risk Assessment Tool (PROBAST)(13). This study aims to bridge this gap through rigorous data collection and preprocessing efforts, alongside the application of sophisticated machine learning methodologies. By establishing a robust, interpretable framework, our research endeavors to empower clinicians with the tools necessary for identifying patients at elevated risk of VTE following a stroke, thereby enabling the formulation of precise interventions to diminish the incidence of this life-threatening complication.

## Materials and methods

### Study Population

The dataset for this investigation was derived from the Shenzhen Neurological Disease System Platform, an extensive repository that has been methodically aggregating detailed data on ischemic stroke patients since 2021. This dataset encompasses a wide range of information, including sociodemographic characteristics, precise details regarding pre-hospital onset, in-hospital diagnostic findings, treatment records, and laboratory test outcomes from 20 affiliated hospitals. The study’s inclusion criteria targeted individuals hospitalized due to stroke between December 2021 and December 2023, who were diagnosed using magnetic resonance imaging (MRI) or computed tomography angiography (CTA), aged 18 years or older, and admitted within seven days of symptom onset as per the International Classification of Diseases, Tenth Revision (ICD-10) criteria. Exclusions were made for patients with transient ischemic attacks, subarachnoid hemorrhage, brain tumors, cerebral venous thrombosis, those diagnosed with distal deep vein thrombosis (DVT) prior to admission, or with a history of pulmonary embolism. For the development and validation of our model, data collected from December 2021 to June 2023 and from July 2023 to December 2023 were utilized, respectively. The platform’s data management and quality control measures were stringently overseen by specialized data managers and quality assurance staff to ensure the anonymization, quality, preservation, and exportation of data met high standards. The data for this research were accessed on January 24, 2024, ensuring all patient data, directly sourced from diagnostic and treatment documentation, were meticulously recorded by experienced neurology specialists for reliability and precision in the dataset used in our research.

### Predictor Variables and Outcomes

The primary objective of this investigation was to construct a predictive model that accurately forecasts the occurrence of VTE following after AIS using a comprehensive dataset representative of a broad spectrum of AIS patient profiles. Patients from the Shenzhen Neurological Disease System Platform, enrolled consecutively from December 2021 to December 2023, formed the basis of this analysis. The selection of potential predictive variables was confined to characteristics documented within the initial three days of hospital admission. These variables, meticulously captured via dedicated case report forms, encompassed a diverse array of factors: gender, ethnicity, age, height, weight, smoking, drinking, diabetes mellitus (DM), hyperlipidemia, atrial fibrillation, history of cerebral infarction, anemia, other comorbid conditions, electrocardiogram (ECG), prehospital medication, pre-morbid mRS, mRS after Admission, NIHSS at onset, NIHSS during hospitalization,glasgow coma scale (GCS), Trial of ORG 10172 in acute stroke treatment(TOAST) classification, weakness, dysarthria, dizziness, paresthesia, headache, convulsion, consciousness status, symptomatic treatment, endovascular treatment (EVT), thrombolytic therapy, lymphocyte count, high-sensitivity c-reactive protein (hsCRP), international normalized ratio (INR), fibrinogen, D-dimer, alanine aminotransferase, low-density lipoprotein cholesterol (LDLC), aspirin, clopidogrel, heparin, enoxaparin, low molecular weight heparin, unfractionated heparin, warfarin, rivaroxaban, sulfonylureas, glycosidase inhibitors, anti-infective treatment, traditional Medicine, lipid medication, anti-platelet therapy, anticoagulant therapy, anti-lipidemic drugs, anti-diabetic yreatment, chinese medicines, and intracranial artery stenosis.

All VTE diagnoses were conclusively determined via color Doppler ultrasound and pulmonary CT angiography (PCTA), serving as the diagnostic benchmarks. The diagnosis of VTE was grounded on a thorough assessment of clinical manifestations coupled with imaging outcomes, primarily focusing on the management of hospitalized patients. A stratified screening methodology was employed for stroke patients manifesting potential symptoms of VTE during their hospital stay, including but not limited to leg pain or swelling, localized warmth, dyspnea, or chest discomfort. This tailored screening approach facilitated the identification and subsequent evaluation of individuals exhibiting clinical presentations suggestive of high-risk VTE, employing color Doppler ultrasound or PCTA for comprehensive examination when warranted.

### Data processing and feature selection

In this study, we harnessed advanced machine learning techniques, notably the K-nearest neighbor (KNN) algorithm and the synthetic minority oversampling technique (SMOTE), to refine our dataset, thereby augmenting the predictive accuracy for VTE risk. This meticulous strategy significantly enhanced the model’s reliability and its capability to generalize across diverse clinical scenarios pertaining to VTE risk assessment. The KNN algorithm was deployed to impute missing values, capitalizing on the proximity of each data point to its nearest "neighbors" within the feature space. This method proved invaluable in preserving the integrity and richness of our dataset, allowing for the retention of complex and multidimensional clinical data without resorting to invasive data collection methodologies. The integrity of the dataset was thus maintained, ensuring minimal loss of information and retaining the nuanced interplay of clinical variables. Regarding the missing number, 30% of the variables were eliminated in this study and were not included in the data analysis(S1 Table). SMOTE addressed the challenge of imbalanced classification inherent in our dataset. By generating synthetic samples for minority classes, it amplified the representation of these underrepresented groups within the dataset. This enhancement was pivotal in improving the model’s sensitivity and accuracy in predicting VTE occurrences, a relatively rare but clinically significant event. The incorporation of SMOTE thus balanced the dataset, ensuring a more equitable representation of all classes and enhancing the model’s predictive precision. Employing these sophisticated data processing and feature selection techniques, our study laid a robust foundation for the development of a highly accurate and generalizable VTE risk prediction model. This approach not only preserved the complexity and diversity of our clinical data but also addressed critical challenges related to data imbalances, setting a new benchmark in the predictive modeling of VTE risks.

### Model development and performance evaluation

In the quest to devise an accurate predictive model for VTE risk following AIS, our study employed a two-pronged approach for feature selection: stepwise forward logistic regression and Least Absolute Shrinkage and Selection Operator (LASSO) analysis. The former method was instrumental in pinpointing variables significantly correlated with VTE risk, whereas LASSO analysis played a critical role in mitigating model overfitting and enhancing variable selection through the imposition of regularization penalties. The construction of the model was underpinned by the application of ten-fold cross-validation, a technique that bolsters both the performance and generalizability of the model. By segmenting the dataset into ten discrete parts, and iteratively training on nine while testing on the remaining one, this procedure guaranteed that each segment was utilized as a test set once, thereby enabling a more nuanced and accurate assessment of the model’s efficacy. To encompass a broad spectrum of analytical perspectives and augment predictive accuracy, the study harnessed an array of machine learning algorithms, including Logistic Regression (LR), Naive Bayes (NB), Decision Trees (DT), Random Forest (RF), Gradient Boosting Machines (GBM), Extreme Gradient Boosting (XGB), and Support Vector Machines (SVM). The distinct attributes and suitability of each algorithm were leveraged to furnish a comprehensive analysis of the data from multifarious angles. Data preprocessing and model building are performed in the python3.9 environment.

### Statistical analysis

Our dataset was subjected to a rigorous descriptive statistical analysis, employing Chi-square tests for categorical variables, T-tests for normally distributed continuous variables, and rank-sum tests for variables deviating from normal distribution. This phase aimed to isolate independent predictors of distal DVT in patients experiencing acute stroke, with variables demonstrating P-values <0.05 advancing to the subsequent feature selection stage. This latter stage utilized both stepwise forward logistic regression and LASSO for variable selection, with the model’s construction synthesizing insights from both methodologies. Visualization techniques, such as scatter plots for illustrating SMOTE sampling outcomes and ROC curve area under the curve (AUC) analysis, provided a graphical representation of cross-validation results and real-world application efficacy for each algorithm. Additionally, SHAP algorithm analysis was employed to enhance the interpretability of model features, particularly for the optimal model. Data analysis is performed in python3.9.

### Ethics approval and consent to participate

This research received the endorsement of the Ethics Review Committee of Shenzhen Longhua District People’s Hospital. In adherence to ethical standards, neurology nurse specialists collected patient data for the Shenzhen Neurological Disease System Platform (SNDSP). The inclusion of patients into the system was contingent upon their ability or willingness to provide written informed consent, either personally or through their representatives, thereby ensuring adherence to ethical research practices.

## Results

### Characteristics of study population

The cohort comprised 1,632 subjects, among which the incidence of VTE was 4.17% (n = 68), with a notable predominance of female patients. The median age of individuals diagnosed with VTE was 69.00 years, significantly older than their non-VTE counterparts, who had a median age of 58.00 years (p < 0.001). Detailed demographic and clinical characteristics of the study population are delineated in Table 1.

**Table 1.**
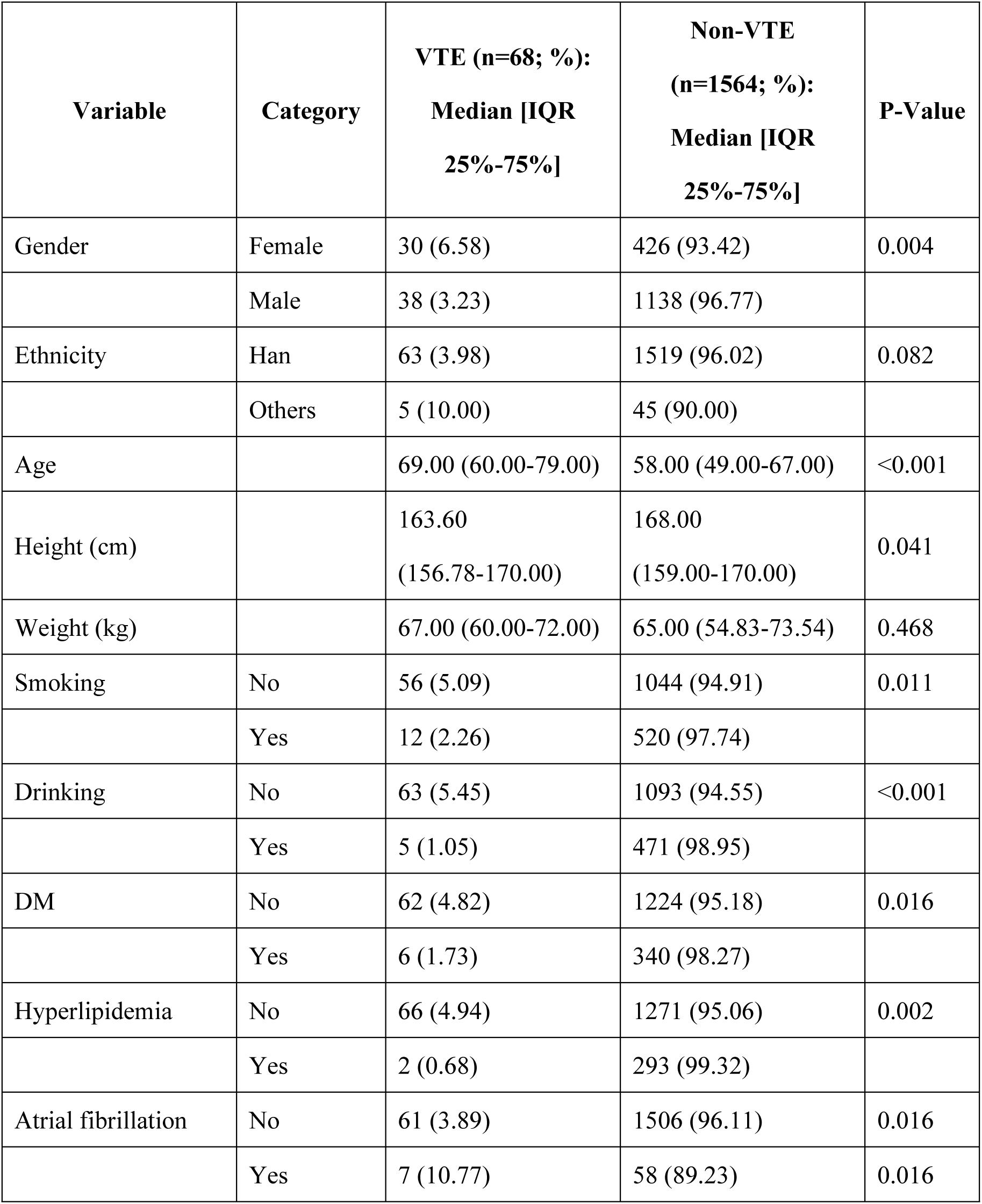
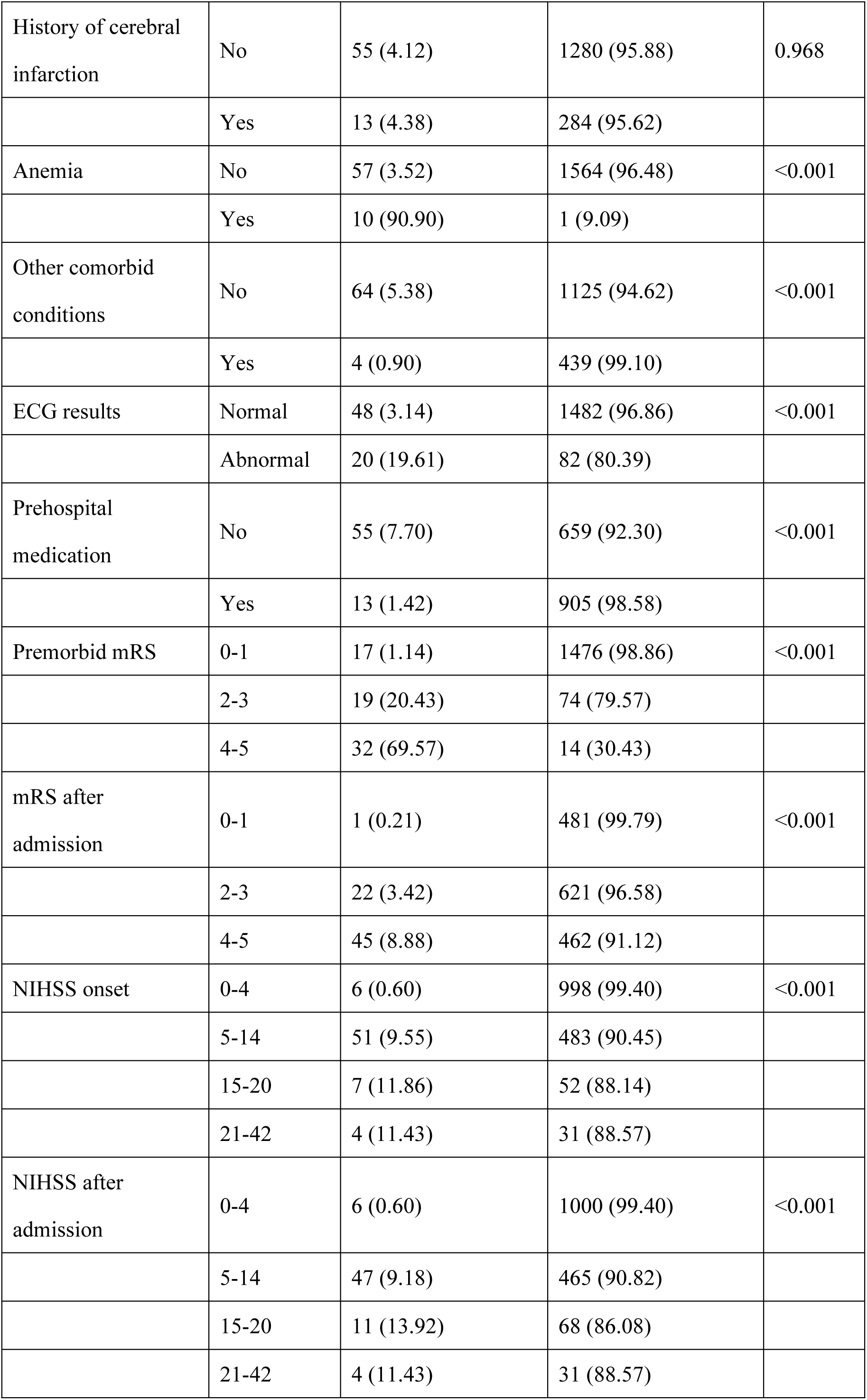
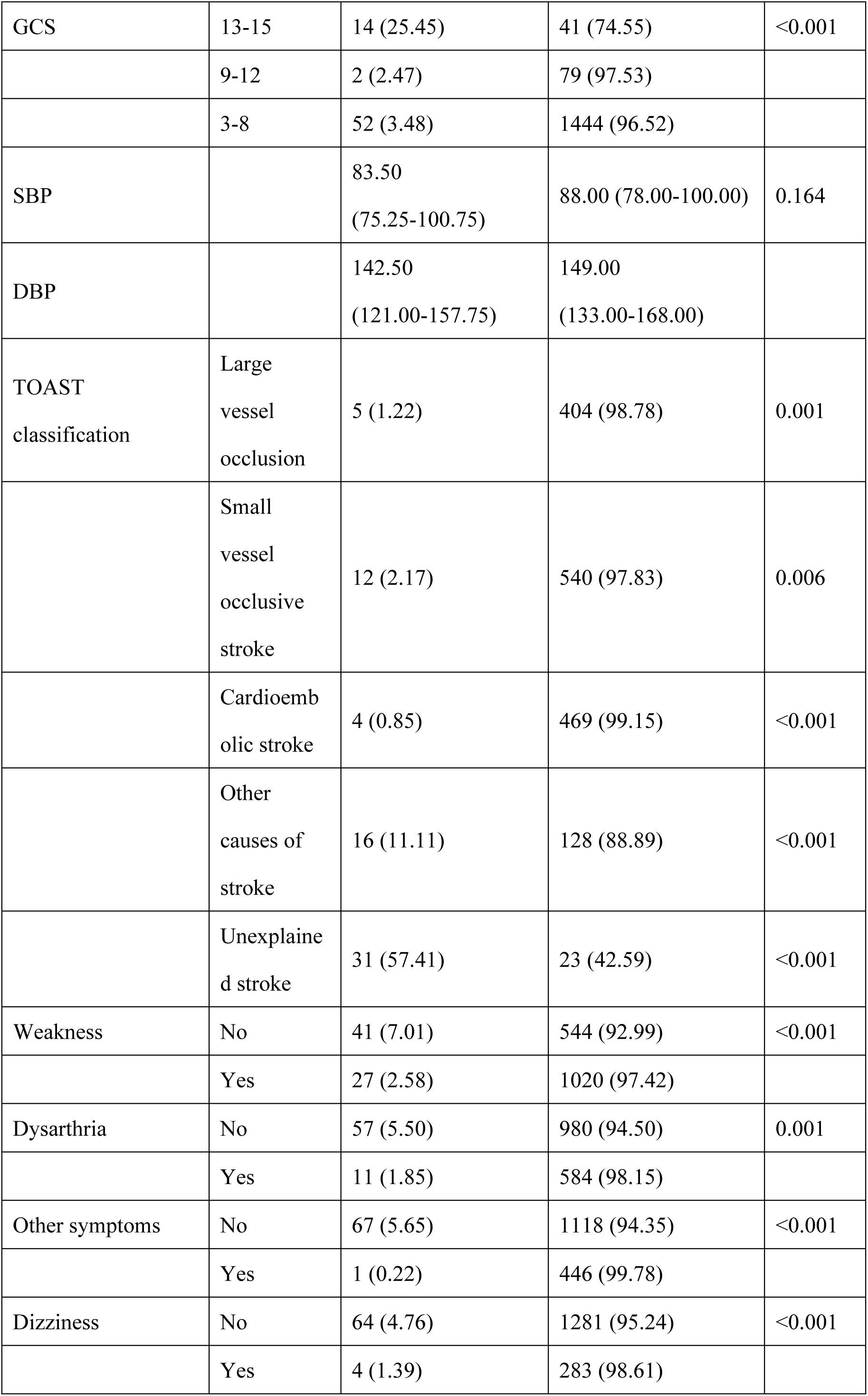
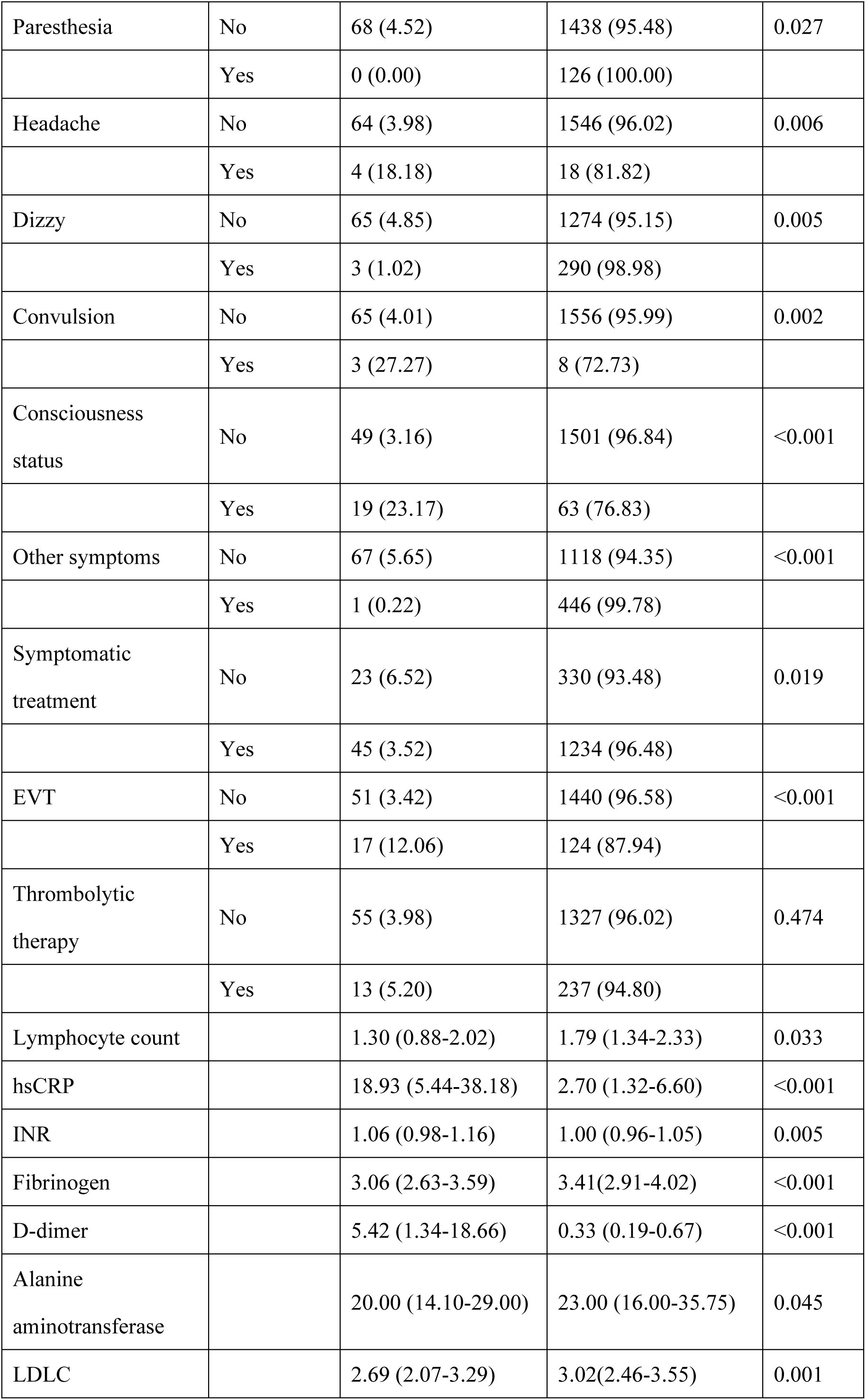
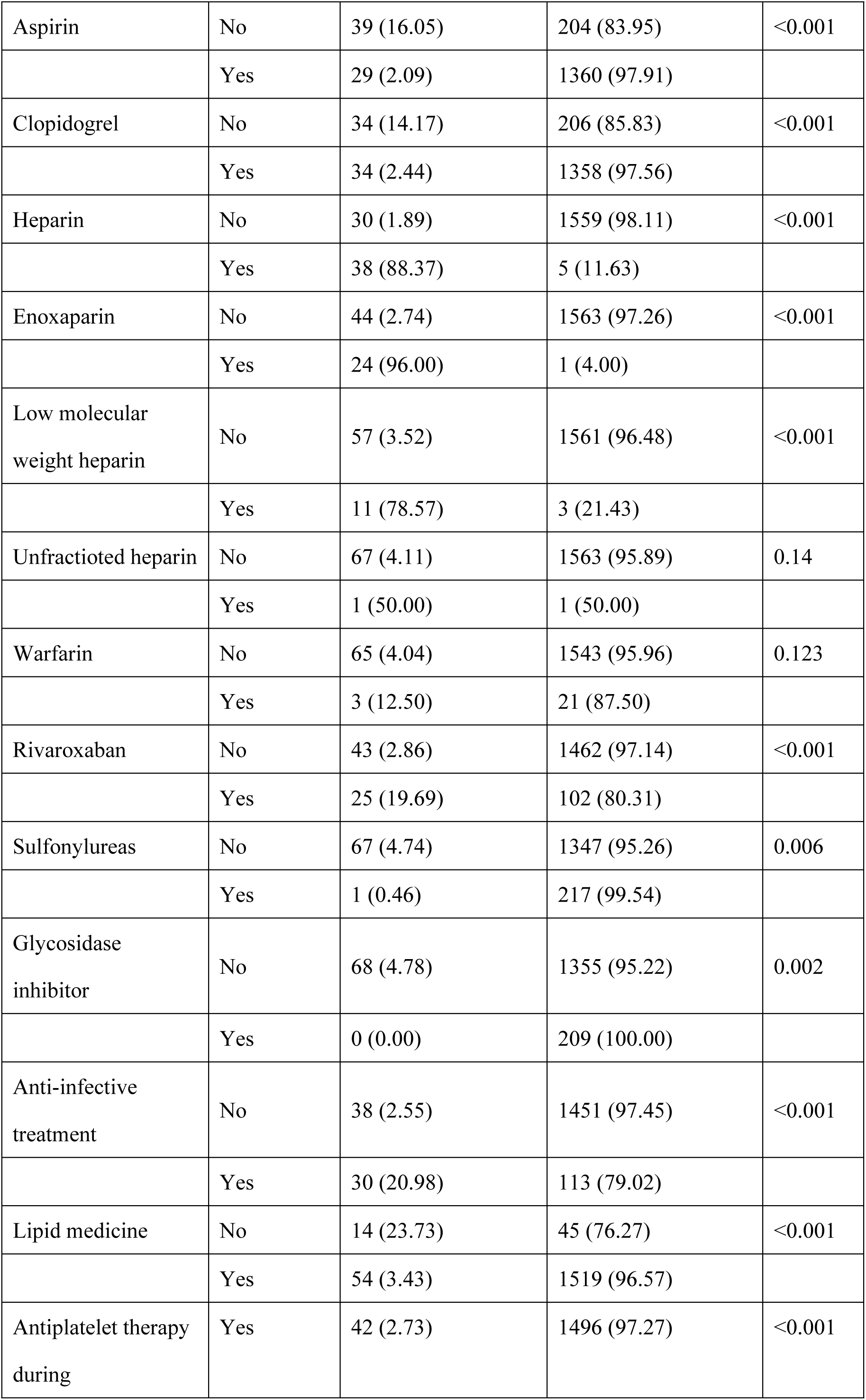
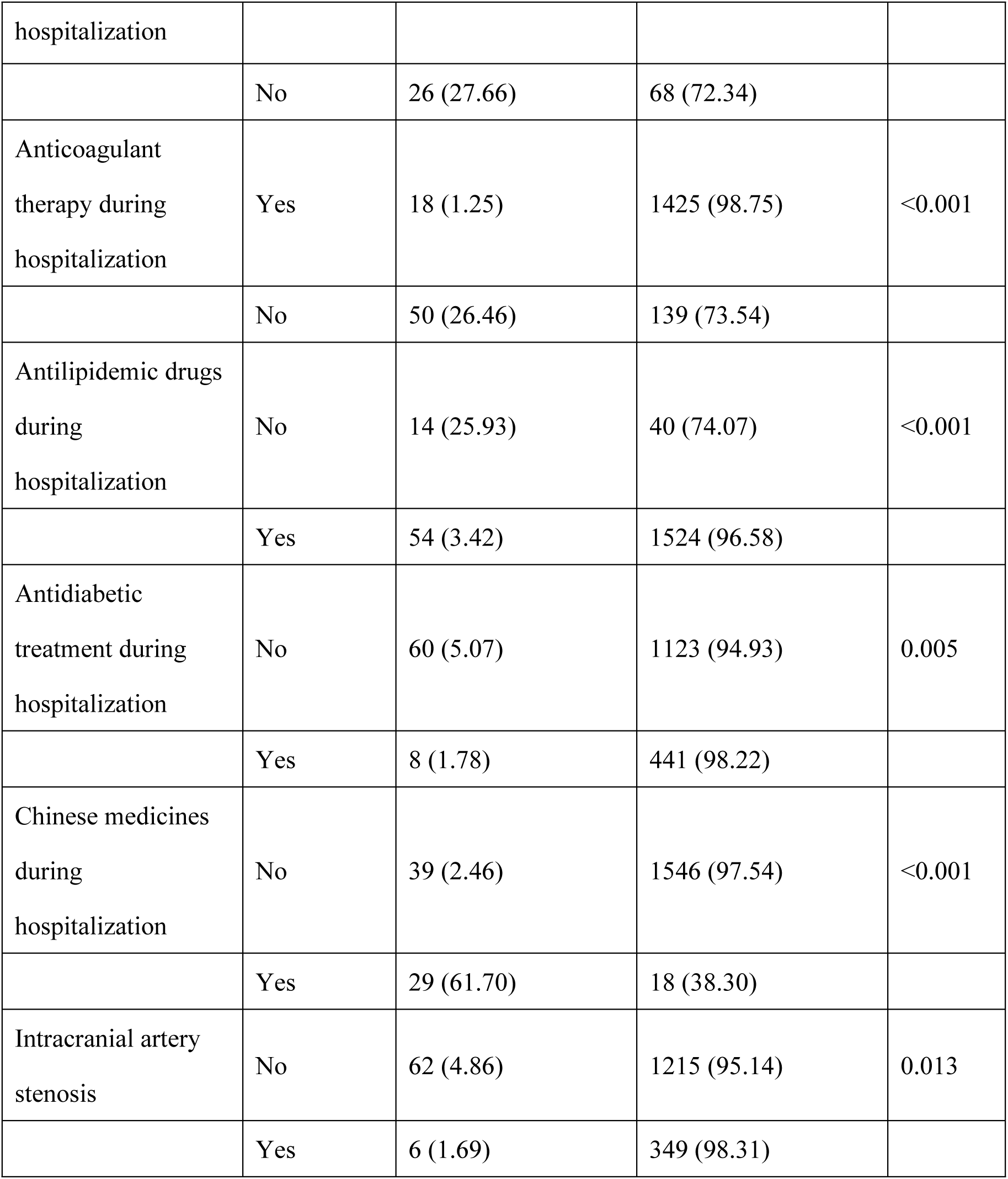
Demographic and Baseline Characteristics by VTE Status.

| Variable | Category | VTE (n=68; %):<br>Median [IQR<br>25%-75%] | Non-VTE<br>(n=1564; %):<br>Median [IQR<br>25%-75%] | P-Value |
| --- | --- | --- | --- | --- |
| Gender | Female | 30 (6.58) | 426 (93.42) | 0.004 |
|  | Male | 38 (3.23) | 1138 (96.77) |  |
| Ethnicity | Han | 63 (3.98) | 1519 (96.02) | 0.082 |
|  | Others | 5 (10.00) | 45 (90.00) |  |
| Age |  | 69.00 (60.00-79.00) | 58.00 (49.00-67.00) | <0.001 |
| Height (cm) |  | 163.60<br>(156.78-170.00) | 168.00<br>(159.00-170.00) | 0.041 |
| Weight (kg) |  | 67.00 (60.00-72.00) | 65.00 (54.83-73.54) | 0.468 |
| Smoking | No | 56 (5.09) | 1044 (94.91) | 0.011 |
|  | Yes | 12 (2.26) | 520 (97.74) |  |
| Drinking | No | 63 (5.45) | 1093 (94.55) | <0.001 |
|  | Yes | 5 (1.05) | 471 (98.95) |  |
| DM | No | 62 (4.82) | 1224 (95.18) | 0.016 |
|  | Yes | 6 (1.73) | 340 (98.27) |  |
| Hyperlipidemia | No | 66 (4.94) | 1271 (95.06) | 0.002 |
|  | Yes | 2 (0.68) | 293 (99.32) |  |
| Atrial fibrillation | No | 61 (3.89) | 1506 (96.11) | 0.016 |
|  | Yes | 7 (10.77) | 58 (89.23) | 0.016 |
| History of cerebral infarction | No | 55 (4.12) | 1280 (95.88) | 0.968 |
|  | Yes | 13 (4.38) | 284 (95.62) |  |
| Anemia | No | 57 (3.52) | 1564 (96.48) | <0.001 |
|  | Yes | 10 (90.90) | 1 (9.09) |  |
| Other comorbid conditions | No | 64 (5.38) | 1125 (94.62) | <0.001 |
|  | Yes | 4 (0.90) | 439 (99.10) |  |
| ECG results | Normal | 48 (3.14) | 1482 (96.86) | <0.001 |
|  | Abnormal | 20 (19.61) | 82 (80.39) |  |
| Prehospital medication | No | 55 (7.70) | 659 (92.30) | <0.001 |
|  | Yes | 13 (1.42) | 905 (98.58) |  |
| Premorbid mRS | 0-1 | 17 (1.14) | 1476 (98.86) | <0.001 |
|  | 2-3 | 19 (20.43) | 74 (79.57) |  |
|  | 4-5 | 32 (69.57) | 14 (30.43) |  |
| mRS after admission | 0-1 | 1 (0.21) | 481 (99.79) | <0.001 |
|  | 2-3 | 22 (3.42) | 621 (96.58) |  |
|  | 4-5 | 45 (8.88) | 462 (91.12) |  |
| NIHSS onset | 0-4 | 6 (0.60) | 998 (99.40) | <0.001 |
|  | 5-14 | 51 (9.55) | 483 (90.45) |  |
|  | 15-20 | 7 (11.86) | 52 (88.14) |  |
|  | 21-42 | 4 (11.43) | 31 (88.57) |  |
| NIHSS after admission | 0-4 | 6 (0.60) | 1000 (99.40) | <0.001 |
|  | 5-14 | 47 (9.18) | 465 (90.82) |  |
|  | 15-20 | 11 (13.92) | 68 (86.08) |  |
|  | 21-42 | 4 (11.43) | 31 (88.57) |  |
| GCS | 13-15 | 14 (25.45) | 41 (74.55) | <0.001 |
|  | 9-12 | 2 (2.47) | 79 (97.53) |  |
|  | 3-8 | 52 (3.48) | 1444 (96.52) |  |
| SBP |  | 83.50<br>(75.25-100.75) | 88.00 (78.00-100.00) | 0.164 |
| DBP |  | 142.50<br>(121.00-157.75) | 149.00<br>(133.00-168.00) |  |
| TOAST<br>classification | Large<br>vessel<br>occlusion | 5 (1.22) | 404 (98.78) | 0.001 |
|  | Small<br>vessel<br>occlusive<br>stroke | 12 (2.17) | 540 (97.83) | 0.006 |
|  | Cardioemb<br>olic stroke | 4 (0.85) | 469 (99.15) | <0.001 |
|  | Other<br>causes of<br>stroke | 16 (11.11) | 128 (88.89) | <0.001 |
|  | Unexplaine<br>d stroke | 31 (57.41) | 23 (42.59) | <0.001 |
| Weakness | No | 41 (7.01) | 544 (92.99) | <0.001 |
|  | Yes | 27 (2.58) | 1020 (97.42) |  |
| Dysarthria | No | 57 (5.50) | 980 (94.50) | 0.001 |
|  | Yes | 11 (1.85) | 584 (98.15) |  |
| Other symptoms | No | 67 (5.65) | 1118 (94.35) | <0.001 |
|  | Yes | 1 (0.22) | 446 (99.78) |  |
| Dizziness | No | 64 (4.76) | 1281 (95.24) | <0.001 |
|  | Yes | 4 (1.39) | 283 (98.61) |  |
| Paresthesia | No | 68 (4.52) | 1438 (95.48) | 0.027 |
|  | Yes | 0 (0.00) | 126 (100.00) |  |
| Headache | No | 64 (3.98) | 1546 (96.02) | 0.006 |
|  | Yes | 4 (18.18) | 18 (81.82) |  |
| Dizzy | No | 65 (4.85) | 1274 (95.15) | 0.005 |
|  | Yes | 3 (1.02) | 290 (98.98) |  |
| Convulsion | No | 65 (4.01) | 1556 (95.99) | 0.002 |
|  | Yes | 3 (27.27) | 8 (72.73) |  |
| Consciousness status | No | 49 (3.16) | 1501 (96.84) | <0.001 |
|  | Yes | 19 (23.17) | 63 (76.83) |  |
| Other symptoms | No | 67 (5.65) | 1118 (94.35) | <0.001 |
|  | Yes | 1 (0.22) | 446 (99.78) |  |
| Symptomatic treatment | No | 23 (6.52) | 330 (93.48) | 0.019 |
|  | Yes | 45 (3.52) | 1234 (96.48) |  |
| EVT | No | 51 (3.42) | 1440 (96.58) | <0.001 |
|  | Yes | 17 (12.06) | 124 (87.94) |  |
| Thrombolytic therapy | No | 55 (3.98) | 1327 (96.02) | 0.474 |
|  | Yes | 13 (5.20) | 237 (94.80) |  |
| Lymphocyte count |  | 1.30 (0.88-2.02) | 1.79 (1.34-2.33) | 0.033 |
| hsCRP |  | 18.93 (5.44-38.18) | 2.70 (1.32-6.60) | <0.001 |
| INR |  | 1.06 (0.98-1.16) | 1.00 (0.96-1.05) | 0.005 |
| Fibrinogen |  | 3.06 (2.63-3.59) | 3.41(2.91-4.02) | <0.001 |
| D-dimer |  | 5.42 (1.34-18.66) | 0.33 (0.19-0.67) | <0.001 |
| Alanine aminotransferase |  | 20.00 (14.10-29.00) | 23.00 (16.00-35.75) | 0.045 |
| LDLC |  | 2.69 (2.07-3.29) | 3.02(2.46-3.55) | 0.001 |
| Aspirin | No | 39 (16.05) | 204 (83.95) | <0.001 |
|  | Yes | 29 (2.09) | 1360 (97.91) |  |
| Clopidogrel | No | 34 (14.17) | 206 (85.83) | <0.001 |
|  | Yes | 34 (2.44) | 1358 (97.56) |  |
| Heparin | No | 30 (1.89) | 1559 (98.11) | <0.001 |
|  | Yes | 38 (88.37) | 5 (11.63) |  |
| Enoxaparin | No | 44 (2.74) | 1563 (97.26) | <0.001 |
|  | Yes | 24 (96.00) | 1 (4.00) |  |
| Low molecular weight heparin | No | 57 (3.52) | 1561 (96.48) | <0.001 |
|  | Yes | 11 (78.57) | 3 (21.43) |  |
| Unfractionated heparin | No | 67 (4.11) | 1563 (95.89) | 0.14 |
|  | Yes | 1 (50.00) | 1 (50.00) |  |
| Warfarin | No | 65 (4.04) | 1543 (95.96) | 0.123 |
|  | Yes | 3 (12.50) | 21 (87.50) |  |
| Rivaroxaban | No | 43 (2.86) | 1462 (97.14) | <0.001 |
|  | Yes | 25 (19.69) | 102 (80.31) |  |
| Sulfonylureas | No | 67 (4.74) | 1347 (95.26) | 0.006 |
|  | Yes | 1 (0.46) | 217 (99.54) |  |
| Glycosidase inhibitor | No | 68 (4.78) | 1355 (95.22) | 0.002 |
|  | Yes | 0 (0.00) | 209 (100.00) |  |
| Anti-infective treatment | No | 38 (2.55) | 1451 (97.45) | <0.001 |
|  | Yes | 30 (20.98) | 113 (79.02) |  |
| Lipid medicine | No | 14 (23.73) | 45 (76.27) | <0.001 |
|  | Yes | 54 (3.43) | 1519 (96.57) |  |
| Antiplatelet therapy during | Yes | 42 (2.73) | 1496 (97.27) | <0.001 |
| hospitalization |  |  |  |  |
|  | No | 26 (27.66) | 68 (72.34) |  |
| Anticoagulant therapy during hospitalization | Yes | 18 (1.25) | 1425 (98.75) | <0.001 |
|  | No | 50 (26.46) | 139 (73.54) |  |
| Antilipidemic drugs during hospitalization | No | 14 (25.93) | 40 (74.07) | <0.001 |
|  | Yes | 54 (3.42) | 1524 (96.58) |  |
| Antidiabetic treatment during hospitalization | No | 60 (5.07) | 1123 (94.93) | 0.005 |
|  | Yes | 8 (1.78) | 441 (98.22) |  |
| Chinese medicines during hospitalization | No | 39 (2.46) | 1546 (97.54) | <0.001 |
|  | Yes | 29 (61.70) | 18 (38.30) |  |
| Intracranial artery stenosis | No | 62 (4.86) | 1215 (95.14) | 0.013 |
|  | Yes | 6 (1.69) | 349 (98.31) |  |

For analytical purposes, the participants were stratified into a training set (n = 1,142) and a test set (n = 490). The composition was predominantly female (71.98% in the training set, 72.24% in the test set) and of Han ethnicity (97.20% in the training set, 96.33% in the test set), with other demographic and clinical attributes showing no significant differences between the two groups, thus ensuring a balanced representation of stroke-related outcomes (Table 2).

**Table 2.** Distribution of Demographic and Clinical Variables in Training and Test Sets.

| **Variable** | **Category** | **Training Set**<br>**(n=1142): %, Median [IQR**<br>**25%-75%]** | **Test Set**<br>**(n=490): %, Median [IQR**<br>**25%-75%]** | **P-Value** |
| --- | --- | --- | --- | --- |
| Gender | Male | 822 (71.98) | 354 (72.24) | 0.960 |
|  | Female | 320 (28.02) | 136 (27.76) | 0.960 |
| Ethnicity | Han | 1110 (97.20) | 472 (96.33) | 0.436 |
|  | Others | 32 (2.80) | 18 (3.67) | 0.436 |
| Age |  | 58.0 (49.0-68.0) | 59.0 (51.0-67.0) | 0.286 |
| Height |  | 168.0<br>(159.0-170.1) | 168.0 (160.0-170.3) | 0.148 |
| Weight |  | 66.93 (59.0-72.0) | 67.32 (60.0-71.922) | 0.293 |
| Smoking | No | 764 (66.90) | 336 (68.57) | 0.547 |
|  | Yes | 378 (33.10) | 154 (31.43) | 0.547 |
| Drinking | No | 806 (70.58) | 350 (71.43) | 0.774 |
|  | Yes | 336 (29.42) | 140 (28.57) | 0.774 |
| DM | No | 904 (79.16) | 382 (77.96) | 0.633 |
|  | Yes | 238 (20.84) | 108 (22.04) | 0.633 |
| Hyperlipidemia | No | 941 (82.40) | 396 (80.82) | 0.489 |
|  | Yes | 201 (17.60) | 94 (19.18) | 0.489 |
| Atrial fibrillation | No | 1101 (96.41) | 466 (95.10) | 0.271 |
|  | Yes | 41 (3.59) | 24 (4.90) | 0.271 |
| History of cerebral infarction | No | 941 (82.40) | 394 (80.41) | 0.376 |
|  | Yes | 201 (17.60) | 96 (19.59) | 0.376 |
| Anemia | No | 1134 (99.30) | 487 (99.39) | 1.000 |
|  | Yes | 8 (0.70) | 3 (0.61) | 1.000 |
| Hyperlipidemia | No | 941 (82.40) | 396 (80.82) | 0.489 |
|  | Yes | 201 (17.60) | 94 (19.18) | 0.489 |
| Other comorbid conditions | No | 819 (71.72) | 370 (75.51) | 0.129 |
|  | Yes | 323 (28.28) | 120 (24.49) | 0.129 |
| ECG | Normal | 1070 (93.70) | 460 (93.88) | 0.978 |
|  | Abnormal | 72 (6.30) | 30 (6.12) | 0.978 |
| Prehospital medication | No | 648 (56.74) | 270 (55.10) | 0.577 |
|  | Yes | 494 (43.26) | 220 (44.90) | 0.577 |
| Pre-morbid mRS | 0-1 | 1048 (91.77) | 445 (90.82) | 0.129 |
|  | 2-3 | 58 (5.08) | 35 (7.14) | 0.129 |
|  | 4-6 | 36 (3.15) | 10 (2.04) | 0.129 |
| mRS after admission | 0-1 | 440 (38.53) | 203 (41.43) | 0.540 |
|  | 2-3 | 361 (31.61) | 146 (29.80) | 0.540 |
|  | 4-6 | 341 (29.86) | 141 (28.78) | 0.540 |
| NIHSS onset | 0-4 | 699 (61.21) | 305 (62.24) | 0.409 |
|  | 5-14 | 372 (32.57) | 162 (33.06) | 0.409 |
|  | 15-20 | 42 (3.68) | 17 (3.47) | 0.409 |
|  | 21-42 | 29 (2.54) | 6 (1.22) | 0.409 |
| NIHSS during hospitalization | 0-4 | 699 (61.21) | 307 (62.65) | 0.282 |
|  | 5-14 | 355 (31.09) | 157 (32.04) | 0.282 |
|  | 15-20 | 59 (5.17) | 20 (4.08) | 0.282 |
|  | 21-42 | 29 (2.54) | 6 (1.22) | 0.282 |
| GCS | 13-15 | 1042 (91.24) | 454 (92.65) | 0.398 |
|  | 9-12 | 57 (4.99) | 24 (4.90) | 0.398 |
|  | 3-8 | 43 (3.77) | 12 (2.45) | 0.398 |
| SBP |  | 88.0 (78.0-100.0) | 87.0 (78.0-99.0) | 0.129 |
| DBP |  |  |  |  |
| TOAST classification | Large vessel occlusion | 288 (25.22) | 121 (24.69) | 0.871 |
|  | Small vessel occlusive stroke | 386 (33.80) | 166 (33.88) | 1.000 |
|  | Cardioembolic stroke | 330 (28.90) | 143 (29.18) | 0.954 |
|  | Other causes of stroke | 96 (8.41) | 48 (9.80) | 0.417 |
|  | Unexplained stroke | 42 (3.68) | 12 (2.45) | 0.262 |
| Weakness | No | 727 (63.66) | 320 (65.31) | 0.562 |
|  | Yes | 415 (36.34) | 170 (34.69) | 0.562 |
| Dysarthria | No | 748 (65.50) | 289 (58.98) | 0.014 |
|  | Yes | 394 (34.50) | 201 (41.02) | 0.014 |
| Other symptoms | No | 820 (71.80) | 365 (74.49) | 0.292 |
|  | Yes | 322 (28.20) | 125 (25.51) | 0.292 |
| Dizziness | No | 944 (82.66) | 405 (82.65) | 1.000 |
|  | Yes | 198 (17.34) | 85 (17.35) | 1.000 |
| Paresthesia | No | 1053 (92.21) | 453 (92.45) | 0.947 |
|  | Yes | 89 (7.79) | 37 (7.55) | 0.947 |
| Headache | No | 1126 (98.60) | 484 (98.78) | 0.961 |
|  | Yes | 16 (1.40) | 6 (1.22) | 0.961 |
| Dizzy | No | 936 (81.96) | 403 (82.24) | 0.947 |
|  | Yes | 206 (18.04) | 87 (17.76) | 0.947 |
| Convulsion | No | 1135 (99.39) | 486 (99.18) | 0.896 |
|  | Yes | 7 (0.61) | 4 (0.82) | 0.896 |
| Other symptoms | No | 820 (71.80) | 365 (74.49) | 0.292 |
|  | Yes | 322 (28.20) | 125 (25.51) | 0.292 |
| Symptomatic treatment | No | 900 (78.81) | 379 (77.35) | 0.554 |
|  | Yes | 242 (21.19) | 111 (22.65) | 0.554 |
| EVT | No | 1040 (91.07) | 451 (92.04) | 0.586 |
|  | Yes | 102 (8.93) | 39 (7.96) | 0.586 |
| Thrombolytic therapy | No | 970 (84.94) | 412 (84.08) | 0.715 |
|  | Yes | 172 (15.06) | 78 (15.92) | 0.715 |
| Lymphocyte count |  | 1.77 (1.32-2.33) | 1.78 (1.34-2.297) | 0.971 |
| hsCRP |  | 2.86 (1.40-7.69) | 2.545 (1.31-6.38) | 0.033 |
| INR |  | 1.01 (0.95-1.06) | 1.00 (0.96-1.06) | 0.507 |
| fibrinogen |  | 3.09 (2.65-3.64) | 3.0385 (2.63-3.52) | 0.189 |
| D-dimer |  | 0.34 (0.19-0.77) | 0.352 (0.21-0.71) | 0.886 |
| alanine<br>aminotransferase |  | 20.0 (14.73-30.0) | 20.0 (15.0-29.0) | 0.754 |
| LDLC |  | 3.0 (2.43-3.55) | 3.01 (2.47-3.52) | 0.860 |
| Aspirin | No | 964 (84.41) | 425 (86.73) | 0.258 |
|  | Yes | 178 (15.59) | 65 (13.27) | 0.258 |
| Clopidogrel | No | 968 (84.76) | 424 (86.53) | 0.397 |
|  | Yes | 174 (15.24) | 66 (13.47) | 0.397 |
| Heparin | No | 1111 (97.29) | 478 (97.55) | 0.890 |
|  | Yes | 31 (2.71) | 12 (2.45) | 0.890 |
| Enoxaparin | No | 1122 (98.25) | 485 (98.98) | 0.378 |
|  | Yes | 20 (1.75) | 5 (1.02) | 0.378 |
| Low molecular weight | No | 1131 (99.04) | 487 (99.39) | 0.680 |
| heparin |  |  |  |  |
|  | Yes | 11 (0.96) | 3 (0.61) | 0.680 |
| Unfractionated heparin | No | 1140 (99.82) | 490.0 (100.00) | 0.877 |
|  | Yes | 2 (0.18) | 0.0 (0.00) | 0.877 |
| Warfarin | No | 1122 (98.25) | 486 (99.18) | 0.225 |
|  | Yes | 20 (1.75) | 4 (0.82) | 0.225 |
| Rivaroxaban | No | 1052 (92.12) | 453 (92.45) | 0.899 |
|  | Yes | 90 (7.88) | 37 (7.55) | 0.899 |
| Sulfonylureas | No | 998 (87.39) | 416 (84.90) | 0.201 |
|  | Yes | 144 (12.61) | 74 (15.10) | 0.201 |
| Glycosidase inhibitor | No | 998 (87.39) | 425 (86.73) | 0.777 |
|  | Yes | 144 (12.61) | 65 (13.27) | 0.777 |
| Anti-infective treatment | No | 1032 (90.37) | 457 (93.27) | 0.072 |
|  | Yes | 110 (9.63) | 33 (6.73) | 0.072 |
| Lipid medicine | No | 1100 (96.32) | 473 (96.53) | 0.951 |
|  | Yes | 42 (3.68) | 17 (3.47) | 0.951 |
| Antiplatelet therapy during hospitalization | No | 1072 (93.87) | 466 (95.10) | 0.388 |
|  | Yes | 70 (6.13) | 24 (4.90) | 0.388 |
| Anticoagulant therapy during hospitalization | No | 1008 (88.27) | 435 (88.78) | 0.833 |
|  | Yes | 134 (11.73) | 55 (11.22) | 0.833 |
| Antilipidemic drugs during hospitalization | No | 1104 (96.67) | 474 (96.73) | 1.000 |
|  | Yes | 38 (3.33) | 16 (3.27) | 1.000 |
| Antidiabetic treatment during hospitalization | No | 832 (72.85) | 351 (71.63) | 0.655 |
|  | Yes | 310 (27.15) | 139 (28.37) | 0.655 |
| Chinese medicines during hospitalization | No | 1103 (96.58) | 482 (98.37) | 0.070 |
|  | Yes | 39 (3.42) | 8 (1.63) | 0.070 |
| Intracranial artery stenosis | No | 1054 (92.29) | 459 (93.67) | 0.380 |
|  | Yes | 88 (7.71) | 31 (6.33) | 0.380 |

### Correlation of variables with clinical outcome

Univariate analysis revealed significant associations between VTE occurrence and several variables, including Gender (p = 0.004), Age (p < 0.001), Height (p = 0.041), Smoking status (p = 0.011), Alcohol consumption (p < 0.001), DM (p = 0.016), Hyperlipidemia (p = 0.002), Atrial fibrillation (p = 0.016), Anemia (p < 0.001), Dysarthria (p = 0.001), ECG findings (p < 0.001), prehospital medication (p < 0.001), pre-morbid mRS (p < 0.001), mRS after admission (p < 0.001), NIHSS at onset (p < 0.001), NIHSS after admission (p < 0.001), GCS (p < 0.001), TOAST classification, weakness, consciousness status, EVT, D-dimer, and LDLC, among other variables. These detailed comparison results are provided in Table 1.

The LASSO model, employing a cross-validation mechanism, fine-tuned the regularization strength (alpha) over a logarithmic scale from 10^-6^ to 10^1^, facilitating precise feature selection. A specified random_state parameter ensured the reproducibility of the findings. The model’s comprehensive analysis underscored the significance of variables such as Pre-morbid mRS, unexplained stroke, in-hospital medications, among others, affirming their relevance to the study’s aims (Fig 1 E and F). Concurrently, stepwise forward logistic regression was utilized to identify pertinent variables for univariate analysis, complementing the LASSO model’s feature selection to guarantee a comprehensive set of predictors for model development. The outcomes of this meticulous variable screening process are presented in S2 Table .

**Figrue 1.**
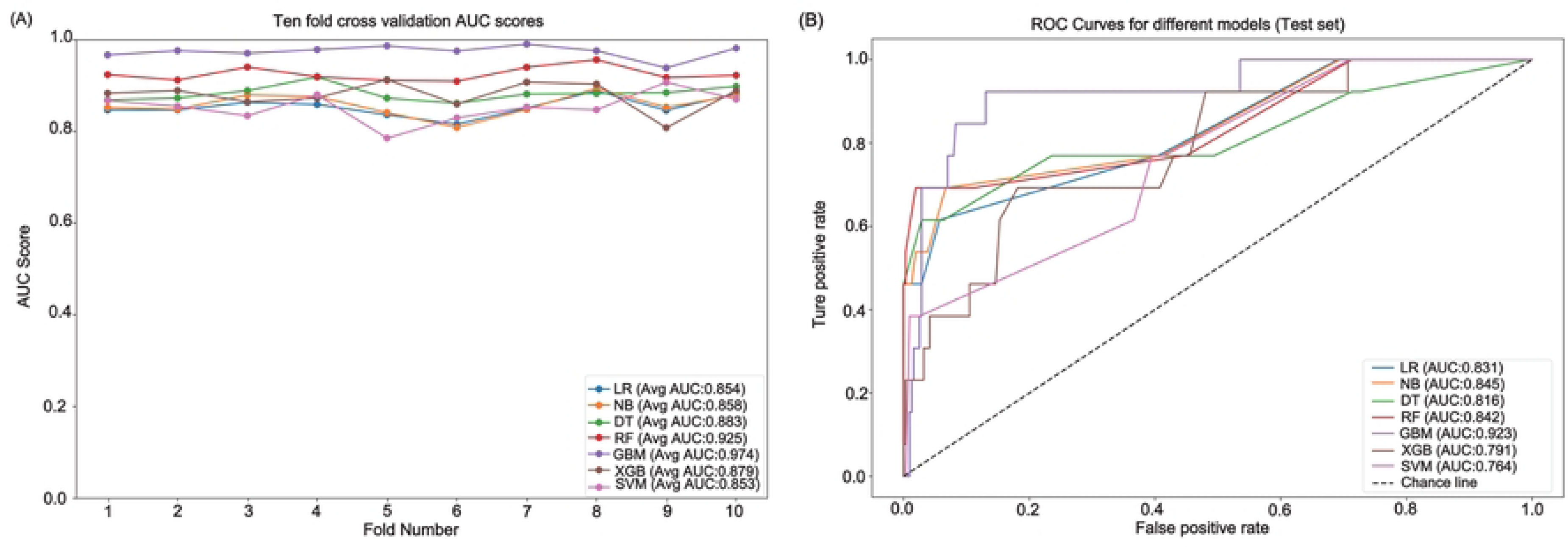
Feature engineering.(A) Original sample visualization using t-SNE shows an imbalanced outcome. (B) Random undersampling addresses imbalanced data by decreasing the majority class. (C) Random oversampling increases the minority class. (D) Synthetic Minority Over-sampling Technique with Nominal Continuous (SMOTE-NC) synthesizes data from the minority class. (E) Lasso Regression Coefficients: Indicates the influence of each feature as determined by the Lasso model. (F) Lasso coefficient path diagram.

### Development and validation of predictive models

Our study employed t-distributed Stochastic Neighbor Embedding (t-SNE) for dimensionality reduction, facilitating a detailed visualization of the distribution patterns between VTE and non-VTE cases within our training dataset (Fig 1). The initial dataset displayed a mixed distribution (Fig 1A), which became sparser following random undersampling (Fig 1B). Conversely, distributions post-oversampling and the application of SMOTE-NC illustrated a more dispersed pattern (Fig 1C and 1D), indicating the significant impact of sampling techniques on data representation.

Through ten-fold cross-validation, we meticulously evaluated the performance of seven distinct machine learning models. The results showcased the Gradient Boosting Machine (GBM) model leading with an AUC score of 0.974, followed by Random Forest (RF) at 0.925, Decision Trees (DT) at 0.883, Extreme Gradient Boosting (XGB) at 0.879, Naive Bayes (NB) at 0.858, Logistic Regression (LR) at 0.854, and Support Vector Machines (SVM) at 0.853 (Fig 2A). Testing these models on an independent set validated their robustness, with five models exhibiting AUC values above 0.8, highlighting the GBM model’s superior predictive accuracy with an AUC of 0.923 (Fig 2B).

**Figure 2.**
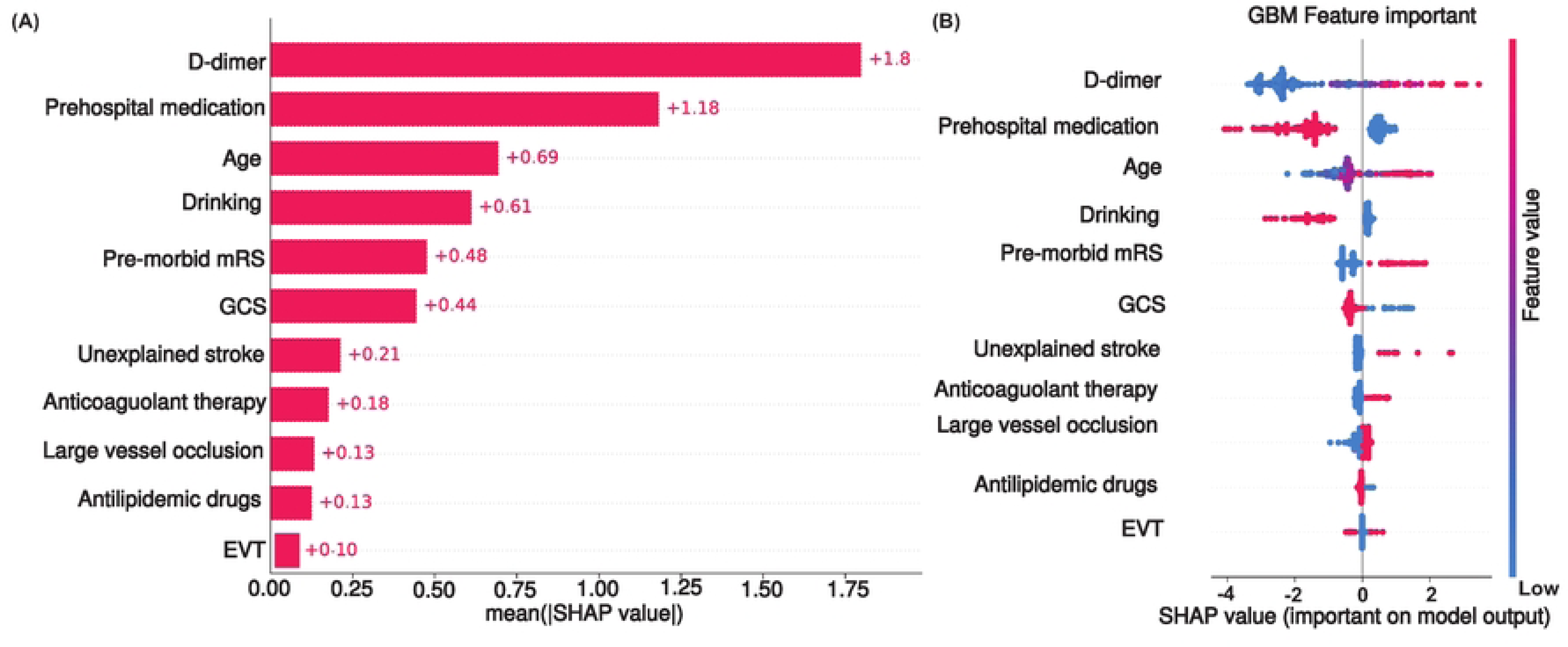
Performance Comparison of Machine Learning Models.(A) Ten-fold cross-validation AUC scores for various models, (B) ROC Curves for different models on the test set.

### Model Interpretation and SHAP Analysis

Ensuring the interpretability of predictive models, particularly in clinical settings, is crucial for their acceptance and application by healthcare professionals. To address this, our study employs SHAP methodology, enabling a transparent evaluation of how each variable influences the model’s predictions. This approach affords a dual-layered interpretation: global insights, which elucidate the model’s overall decision-making process, and local insights, which provide individualized explanations. The global interpretative framework is visualized through SHAP summary plots (Fig 3A and B), where the mean SHAP values of each feature are calculated and ranked. This hierarchy underscores the relative importance of predictors such as D-dimer levels, Prehospital medication, and Age, among others, in determining VTE risk. SHAP dependency plots further dissect the relationship between specific features and the prediction outcome, offering a granular understanding of feature impact.

**Figure 3.**
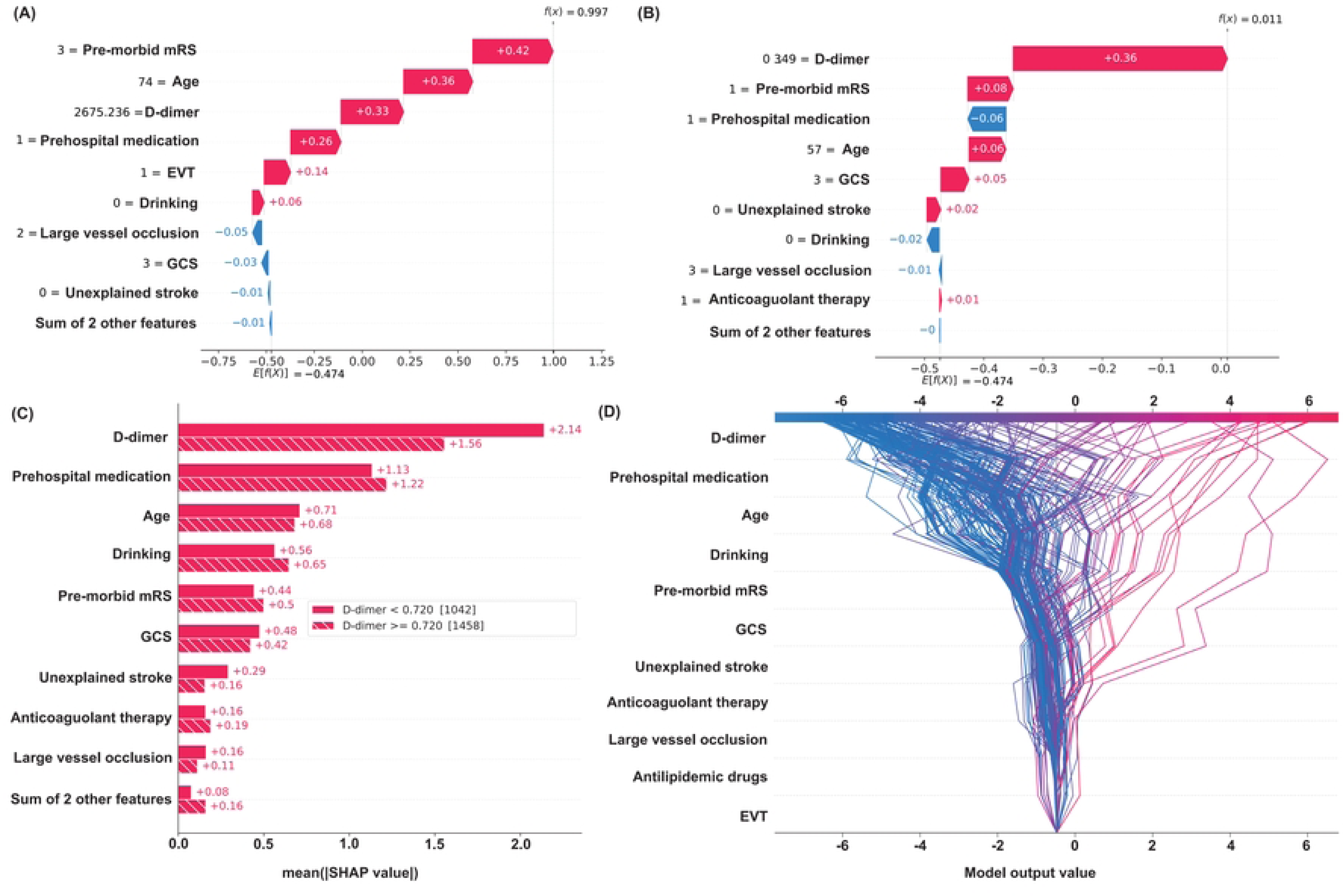
Feature Impact Analysis.(A) Mean SHAP values for predictors. (B) SHAP value distribution for GBM.

Personalized risk assessments, as demonstrated in Fig 4A and B, highlight the model’s ability to integrate individual patient data to predict VTE risk accurately. For instance, one patient was identified with a 99.7% VTE risk, with significant factors being premorbid mRS and age. Conversely, another patient presented a low risk of 1.1%, with premorbid mRS contributing negatively to VTE risk, indicating how diverse variables can influence individual risk profiles differently. Such analyses enable tailored patient care and informed risk management.

**Figure 4.**
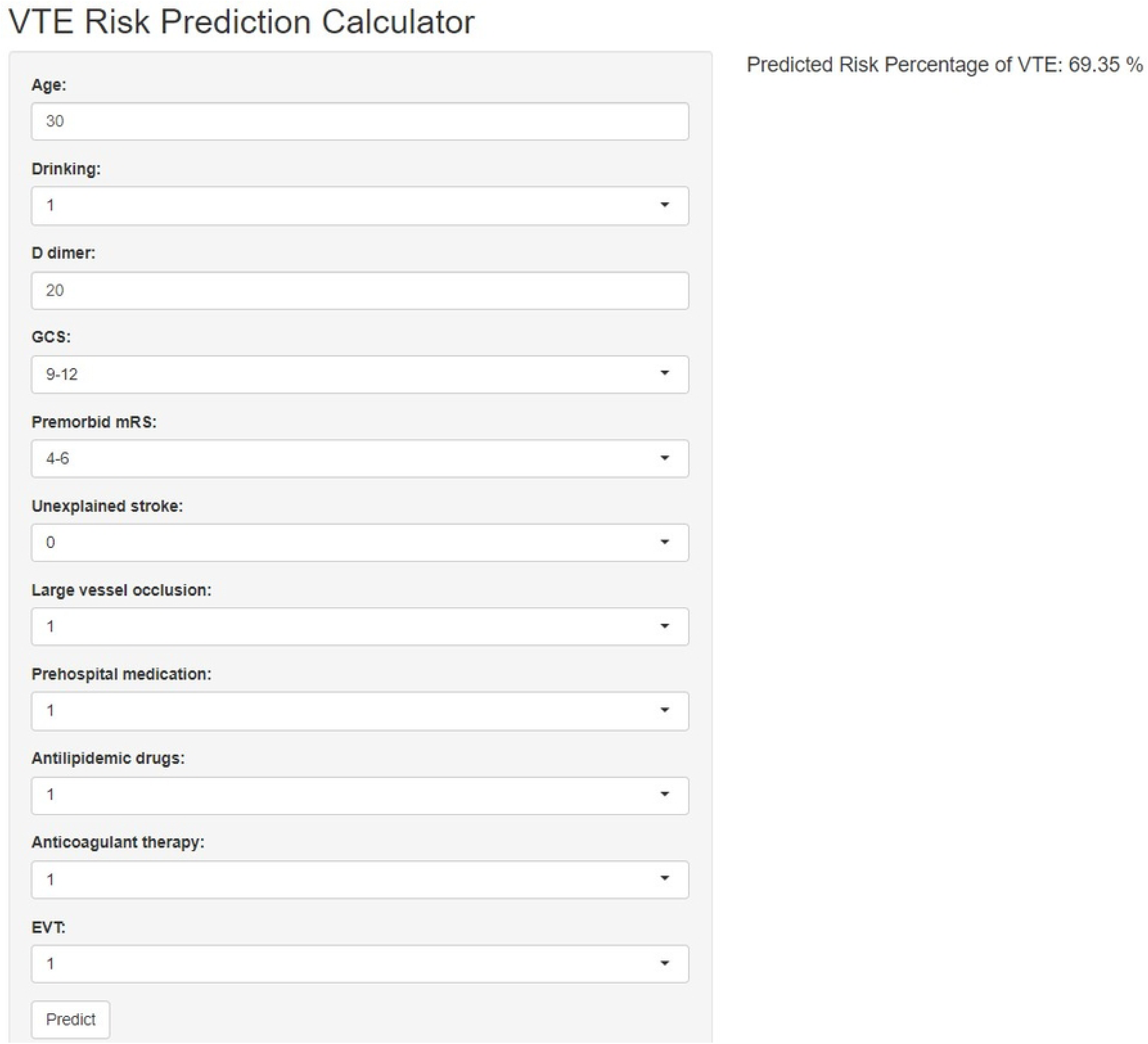
SHAP Value Analysis for Model Prediction.(A) Average impact on model output. (B) Impact of top features on a single prediction. (C) Aggregate SHAP values. (D) SHAP decision plot.

Furthermore, Fig 4C reveals a nonlinear association between D-dimer levels and VTE risk, pinpointing a threshold beyond which VTE risk escalates significantly. This insight is critical for identifying patients who might benefit from closer monitoring or preventive interventions. The SHAP dependency graph (Fig 4D) elaborates on the effect of individual variables across the patient cohort, providing a comprehensive overview of the model’s predictive dynamics.

### Prognostic implications

The exemplary performance of the GBM model culminated in its integration into a user-friendly web application, designed to predict VTE risk in AIS patients based on the model’s key variables. This digital tool, accessible at https://youlijiang236.shinyapps.io/myapp/, empowers clinicians to leverage our predictive model in real-time, facilitating personalized patient care and informed risk management strategies (Fig 5).

**Figure 5.**
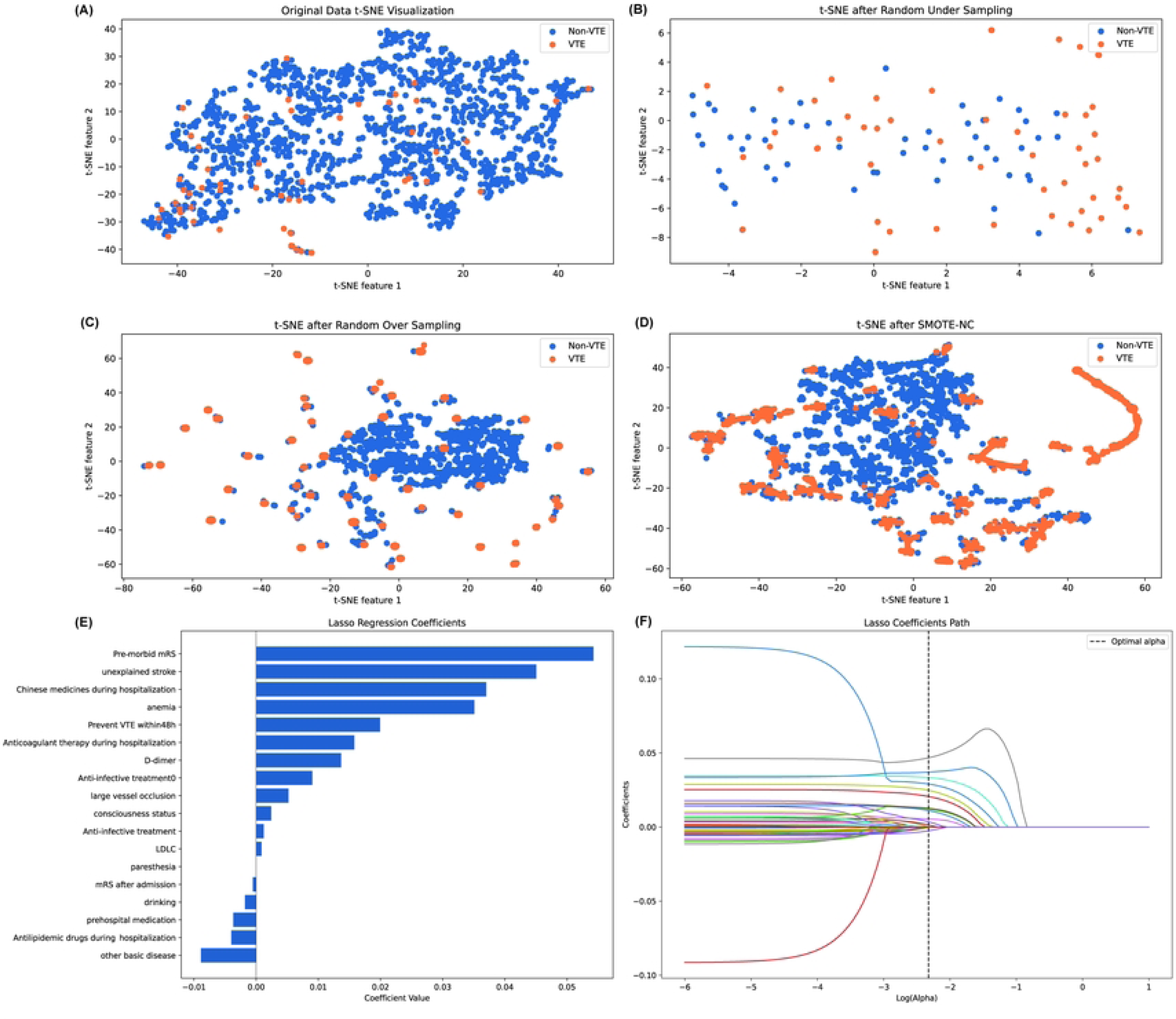
VTE Risk Assessment Tool.

## Discussion

Our investigation has culminated in the development of an array of machine learning models adept at predicting VTE risk among patients suffering from AIS. A distinguishing feature of our models lies in their capacity to amalgamate an extensive array of predictive variables, spanning from elementary demographic information to intricate clinical observations and laboratory findings. This comprehensive approach substantially surpasses the precision and predictive power of conventional risk assessment methodologies(14, 15). Crucially, our models incorporate a broad spectrum of demographic, clinical, and laboratory parameters, thereby offering a holistic risk evaluation framework. Unique to our research is the inclusion of variables previously overlooked in model studies, such as EVT, alongside other variables that reflect the routine diagnostic and therapeutic practices in stroke management. This ensures the practicality and applicability of our model in real-world clinical settings. The employment of the gradient boosting machine (GBM) algorithm marks a significant enhancement over existing predictive approaches(16), not merely in terms of predictive accuracy but also in handling voluminous and readily available variables. A pivotal aspect of our model is its interpretability, facilitated by SHAP value analysis, which demystifies the impact of each predictor on VTE risk. This interpretability equips clinicians with a profound understanding of the prediction process, thereby empowering them to make well-informed and precise clinical decisions. Our model improves the accuracy of VTE risk prediction and enriches the transparency and understandability of prediction results. This dual advantage heralds a novel era in the management of VTE risk post-stroke, spotlighting the potential of machine learning in transforming clinical decision-making and patient care strategies. A review of extant literature reveals a predominant reliance on various forms of logistic regression models for constructing VTE risk prediction frameworks. Notable examples include the DVT risk model developed by Pan et al. using data from patients with acute stroke, which reported a final AUC of 0.785(17), and the post-AIS model by Bonkhoff et al., utilizing L1 regular logit, achieving an AUC of 0.730(18). These studies underscore the conventional approach towards VTE risk prediction, often limited by the methodologies employed.In stark contrast, our study introduces a machine learning model that exhibits superior predictive performance, as evidenced by a notably higher AUC, particularly with the implementation of the GBM algorithm, which attained an AUC of 0.923. This significant improvement in predictive accuracy is further augmented by our innovative approach to feature selection, combining LASSO with stepwise logistic regression. This methodological synergy not only enhances variable selection accuracy and model stability but also considerably reduces the risk of overfitting, thereby bolstering the model’s generalizability—a critical advancement corroborated by multiple preceding studies(19). Furthermore, our meticulous adherence to PROBAST (Predictive Model Biomarker Assessment Tool) standards throughout the processes of data collection, preprocessing, and modeling underscores our commitment to bridging the existing gaps in the literature. Our objective was to construct a model of heightened accuracy capable of identifying patients at elevated risk of VTE more effectively. This ambition was rooted in the recognition of the limitations inherent in previous studies and driven by the potential of advanced machine learning techniques to revolutionize the predictive modeling landscape in VTE risk assessment following acute ischemic stroke.

Our study’s integration of SHAP analysis has illuminated the critical roles of individual risk factors in the genesis of VTE. Among these, prominent factors such as heightened D-dimer levels, prior medication use, patient age, pre-morbid mRS, GCS, the presence of large vessel occlusion, and specific medical interventions emerged as significant predictors. These findings resonate with varying degrees of corroboration from prior research endeavors, underscoring their empirical validity(20).

Notably, our analysis underscored the pivotal biomarker role of D-dimer in forecasting VTE risk, establishing an early warning threshold at a D-dimer level of 0.72 ug/ml. The elevation of D-dimer, a fibrin degradation product indicative of thrombosis and fibrinolytic activity, has been validated in earlier studies as a crucial diagnostic marker for thrombotic events(21, 22).

Concurrently, the GCS score, which gauges a patient’s consciousness level, has been linked to VTE risk, with lower scores suggesting compromised brain function and consequently an elevated VTE risk(23). Furthermore, our analysis corroborates the heightened VTE susceptibility among older patients and those with diminished pre-stroke functional capacity, as indicated by their premorbid mRS scores(24, 25). Such susceptibility is attributed to diminished mobility and circulation, factors that significantly contribute to thrombosis risk in this patient cohort. Prehospital medications, encompassing antidiabetic and antihypertensive drugs, antibiotics, and traditional Chinese medicine, imply pre-existing health conditions that, coupled with AIS-induced mobility constraints, predispose patients to thrombotic complications(26). The administration of antiplatelet and anticoagulant therapies during hospitalization not only serves as a preventive strategy against thrombotic events but also signals a higher baseline embolic risk. Moreover, the use of antilipidemic medications underscores an indirect VTE risk associated with cardiovascular diseases, highlighting the intricate interplay between cardiovascular health and VTE incidence(27).

Furthermore, our findings indicate that stroke patients who consume alcohol are at an elevated risk of developing post-stroke VTE compared to those who do not drink. This heightened risk may be attributable to alcohol’s indirect influence on the coagulation and fibrinolytic systems, disrupting the delicate balance necessary for normal blood clotting and dissolution(28). The nuanced effects of different treatments on VTE risk during the acute phase of AIS patients have been somewhat overlooked in previous research(29). Nonetheless, our analysis reveals that patients undergoing EVT may face a greater VTE risk compared to those receiving thrombolytic therapy or symptomatic care alone. This increased risk could stem from the prolonged immobilization and delayed return to daily activities post-EVT, as EVT, despite its efficacy in managing acute large vessel occlusion strokes, necessitates extended bed rest(30). This shows that post-operative prevention of stroke patients receiving EVT should pay more attention to the prevention of VTE and strengthen the accessibility of VTE monitoring and examination after EVT.

By amalgamating advanced machine learning techniques with SHAP value analysis, we have developed a server-based web calculator. This innovative tool is poised to revolutionize VTE risk screening post-AIS by enabling the input of essential patient characteristics into the model. It equips healthcare professionals with the capability to precisely identify patients at elevated risk of VTE, thereby facilitating the formulation of targeted prevention and intervention strategies grounded in a meticulous evaluation of risk factors.For instance, the early identification of patients exhibiting high D-dimer levels and diminished GCS allows for the swift adoption of more aggressive preventive approaches, such as anticoagulant therapy, alongside enhanced physical assessments and nursing care, especially for patients receiving EVT. Furthermore, the interpretability afforded by the model’s SHAP values grants physicians deeper insights into the influence of each predictor on VTE risk. This enhanced understanding enables a more nuanced consideration of various treatment modalities’ benefits and drawbacks, guiding clinical decision-making towards optimized antithrombotic treatment strategies and personalized patient care plans.

## Conclusion

Our study advances the prediction of VTE risk in patients with acute ischemic stroke through the application of the GBM algorithm. This approach offers a refined risk assessment tool that can significantly enhance early detection and management of VTE in this vulnerable population. The incorporation of SHAP values for interpretability strengthens the model’s applicability in clinical decision-making, allowing for the development of tailored treatment plans. The potential integration of this model into clinical decision support systems represents a promising direction for future research, aiming to improve clinical efficiency and patient outcomes in a practical healthcare setting.

## Data Availability

All data files related to this study can be obtained from the inquiry (Qingshi Zhao).

## Acknowledgments

We would like to thank the doctors and nurses at the Department of Neurology, Longhua District People’s Hospital, Shenzhen for their help and support in this study.

